# Transport effect of COVID-19 pandemic in France

**DOI:** 10.1101/2020.07.27.20161430

**Authors:** Lina Guan, Christophe Prieur, Liguo Zhang, Clémentine Prieur, Didier Georges, Pascal Bellemain

## Abstract

An extension of the classical pandemic SIRD model is considered for the regional spread of COVID-19 in France under lockdown strategies. This compartment model divides the infected and the recovered individuals into undetected and detected compartments respectively. By fitting the extended model to the real detected data during the lockdown, an optimization algorithm is used to derive the optimal parameters, the initial condition and the epidemics start date of regions in France. Considering all the age classes together, a network model of the pandemic transport between regions in France is presented on the basis of the regional extended model and is simulated to reveal the transport effect of COVID-19 pandemic after lockdown. Using the the measured values of displacement of people mobilizing between each city, the pandemic network of all cities in France is simulated by using the same model and method as the pandemic network of regions. Finally, a discussion on an integro-differential equation is given and a new model for the network pandemic model of each age class is provided.

## 1 Introduction

Up to now, COVID-19 has widely spread over the world and is much more contagious than expected. The outbreak of COVID-19 has resulted in a huge pressure of hospital capacity and a massive death of population in the world. Quarantine and lockdown measures have been taken in many countries to control the spread of the infection, and has proved the amazingly effectiveness of these measures for the outbreak of COVID-19, in particular in China (see [1]). Quarantine is a rather old technique to prevent the spread of diseases. It is used at the individual level to constrain the movement of all the population and encourage them stay at home. Lockdown measures reduce the pandemic transmission by increasing social distance and limiting the contacts and mobility of people, e.g. with cancellation of public gatherings, the closure of public transportation, the closure of borders. COVID-19 may yield a very large number of asymptomatic infected individuals, as mentioned in [2] and [3]. Therefore, most countries have implemented indiscriminate lockdown. But the long time of duration of lockdown can cause inestimable financial costs, many job losses, and particularly psychological panic of people and social instability of some countries.

As declared by some governments (see [4]), testing is crucial to exit lockdown, mitigate the health harm and decrease the economic expensation. In this paper, we consider two classes of active detection. The first one is the short range test: molecular or Polymerase Chain Reaction (PCR) test, that is used to detect whether one person has been infected in the past. The second test is the long range test: serology or immunity test, that allows to determine whether one person is immune to COVID-19 now. This test is used to identify the individuals that cannot be infected again.

For our research on COVID-19, we aim to evaluate the effect of lockdown within a given geographical scale in France, such as the largest cities, or urban agglomerations, or French departments, or one of the 13 Metropolitan Regions (to go from the finest geographical scale to the largest one). The estimations of effect are also considered on different age-classes, such as early childhood, scholar childhood, working class groups, or the elderly. Besides, we propose to understand the effect of partial lockdown or other confinement strategies depending on some geographical perimeters or some age groups (as the one that Lyon experienced very recently, see [5])

In the context of COVID-19, there have been many papers that focus on estimating the effect of lockdown strategies on the spread of the pandemic (e.g. [6] and [7]). In [8], the lockdown effect is estimated using stochastic approximation, expectation maximization and an estimation of basic reproductive numbers. In this work, we aim at evaluating the dynamics of the pandemic after the lockdown by looking on the transport effect.

In this paper, one contribution is that an extension of the typical SIRD pandemic model is presented for characterizing the regional spread of COVID-19 in France before and after the lockdown strategies. Taking into account the detection ratios of infected and immune persons, this extended compartment model integrates all the related features of the transmission of COVID-19 in the regional level. In order to estimate the effect of lockdown strategies and understand the evolution of the undetected compartments for each region in France, an optimization algorithm is used to derive the optimal parameters for regions by fitting the extended model to real reported data during the lockdown.

Based on regional model analysis before and after the lockdown, we present a network model to characterize the pandemic transmission between regions in France after lockdown and evaluate the transport effect of COVID-19 pandemic, when considering all age classes together. The most interesting point is the chosen exponential transmission rate function *β*(*t*), in order to incorporate the complex effect of lockdown and unlockdown strategies and the delay of incubation.

This paper is organized as follows. In Section 2, the extended model is derived from the classical pandemic SIRD model and the rationale behind the model is explained. In Section 3, we present the parameters optimization problem and estimate the effect of lockdown strategies. From the calibration of parameters for each region in France, we derive the pandemic start date of regions. Network model of pandemic transmission between regions is introduced and the network simulation is implemented in Section 4. In Section 5, using the same model as the pandemic network of regions in France, we simulate the pandemic network of all cities in France. In the ‘Discussion’ section, considering the age classification, an integro-differential model is presented for the pandemic network transmission, at any geographical scale, and for any set of age classes.

## 2. Pandemic Model

In this paper, the scenario we consider is a large safe population into which a low level of infectious agent is introduced and a closed population with neither birth, nor natural death, nor migration. There is one basic model of modelling pandemic transmission which is well known as susceptible-infected-recovered-dead (SIRD) model in [9]. This mathematical compartmental model is described as follows,

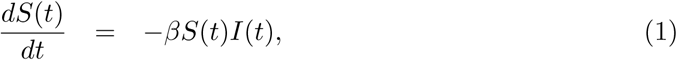

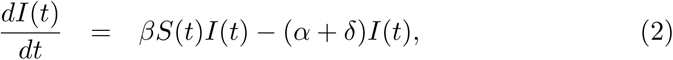

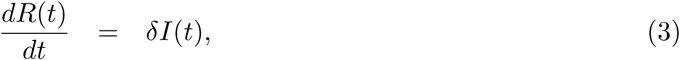

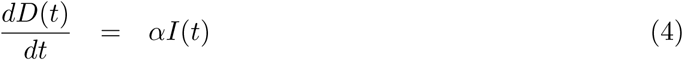

where *S*(*t*) is the number of susceptible people at time *t, I*(*t*) is the number of infected people at time *t, R*(*t*) is the number of recovered people at time *t, D*(*t*) is the number of deaths due to pandemic until time *t*, with constant parameters: *β* is transmission rate per infected, *δ* is the removal or recovery rate, *α* is the disease mortality rate. The compartment variables *S*(*t*), *I*(*t*), *R*(*t*), *D*(*t*) satisfy

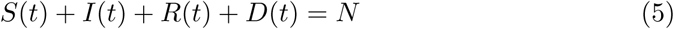

at any time instant *t*, here *N* is the total number of population of the considered area.

From the differential equations (1)-(4), it is obvious that at any time instant *t*, the total rate *βI*(*t*) of transmission from entire susceptible compartment to infected compartment is proportional to the infected *I*; the infected individuals recover at a constant rate *δ*; the infected go to death compartment at a constant rate *α*.

In fact, with the exception of the detected well-known data, there are some undetected data that cannot be measured but are significantly important for the analysis of the evolution of COVID-19 in France under lockdown policy. Moreover they are useful to provide efficient social policies, such as optimal management of limited healthcare resources, the ideal decision of the duration and level of lockdown or re-lockdown, and so on.

Inspired by [10], the basic SIRD model is extended to a more sophisticated compartmental model which includes several features of the recent COVID-19 outbreak, with flexibility with respect to lockdown and test strategies. More sophisticated models could be considered, however it is important that the model we consider can be calibrated with the available data for French regions. On the basis of *SIDUHR*^+/−^ model in [10], this model additionally considers that the infected undetected individuals *I*^−^ and the infected detected individuals *I*^+^ get sicker and then go to intensive care *U*, and the hospitalized individuals *H* die (*D*) before attaining intensive care *U*. In our model, the short term tests transfer the positive individuals from compartment *I*^−^ to compartment *I*^+^. The detection using antibody tests allows to transfer individuals from compartment *R*^−^ to compartment *R*^+^. The presence of antibodies indicate that one person has recovered from the pandemic and is immune. The flow diagram of this model is sketched out in Figure 1.

**Figure 1:**
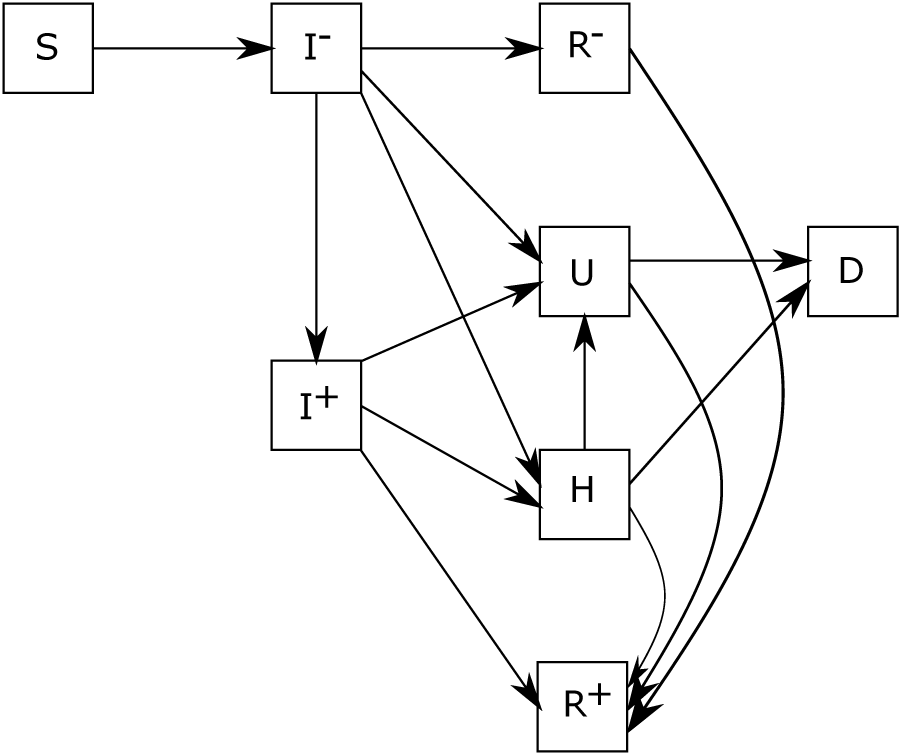
Compartments and flow of the pandemic model (6)-(13).

The evolution of each compartments is modelled by the following equations,

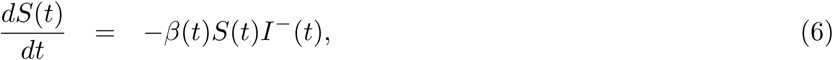

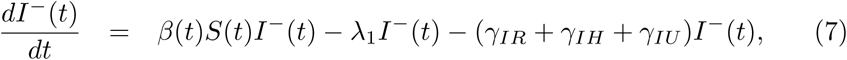

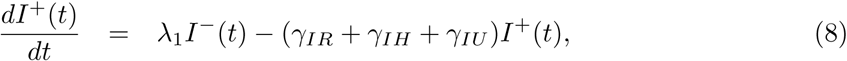

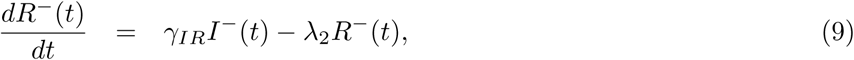

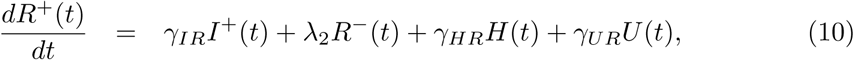

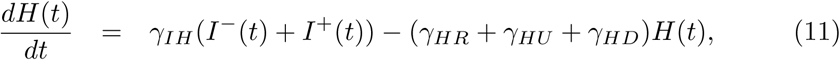

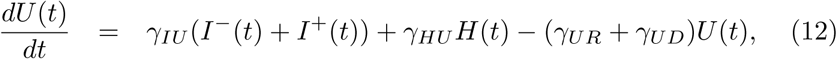

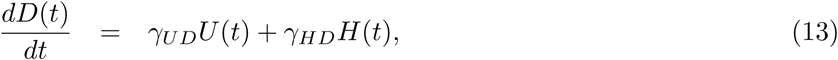

With

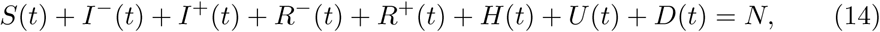

and initial conditions

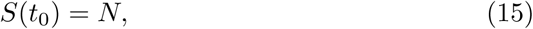

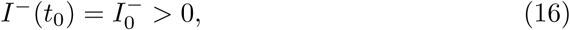

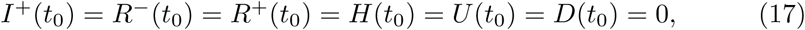

where *S*(*t*) is the number of susceptible individuals at time *t, I*^−^(*t*) is the number of infected undetected individuals at time *t, I*^+^(*t*) is the number of infected detected individuals at time *t, R*^−^(*t*) is the number of recovered undetected individuals at time *t, R*^+^(*t*) is the number of recovered detected individuals at time *t, H*(*t*) is the number of hospitalized individuals at time *t, U* (*t*) is the number of individuals hospitalized in an intensive care unit at time *t, D*(*t*) is the cumulative number of dead individuals from hospital or intensive care at time *t*. We do not consider here deaths from nursing homes for example, as in [11, Chapter 6], where a slightly different is considered at the French national scale. The main reason for that is the lack of data. Indeed, daily data on the total reported cases are unavailable in France at the regional scale. The initial conditions (15)-(17) means that infected people 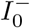 are introduced into a population consisting of susceptible individuals *S*(*t*_0_) at time instant *t*_0_. Both 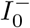 and *t*_0_ are two unknown parameters that need to be identified.

Two types of tests are taken into account in this model, one is a class of virological tests like nasal ones that can detect new infectious cases from compartment *I*^−^. The rate of these tests is denoted by *λ*_1_; another method is a class of serological tests that detect the individuals of infected and sequentially recovered from compartment *R*^−^ applying blood or saliva samples, the rate of these tests (for example, blood test) is denoted by *λ*_2_. This second type of tests was not proposed in France until very recently, thus we consider in this work that *λ*_2_ = 0.

The other parameters in equation (6)-(13) are defined as follows:

- *γ*_*IR*_ is the daily individual transition rate from *I* to *R*, and *γ*_*IR*_ = (1 − *p*_*a*_)(1 − *p*_*H*_)/*N*_*s*_ + *p*_*a*_/*N*_*a*_,
- *γ*_*IH*_ is the daily individual transition rate from *I* to *H*, and *γ*_*IH*_ = (1 − *p*_*a*_)*p*_*H*_ (1 − *p*_*U*_)/*N*_*IH*_,
- *γ*_*IU*_ is the daily individual transition rate from *I* to *U*, and *γ*_*IU*_ = (1 − *p*_*a*_)*p*_*H*_*p*_*U*_ /*N*_*IH*_,
- *γ*_*HR*_ is the daily individual transition rate from *H* to *R*, and *γ*_*HR*_ = (1 − *p*_*HD*_)/*N*_*HR*_,
- *γ*_*HD*_ is the daily individual transition rate from *H* to *D*, and *γ*_*HD*_ = *p*_*HD*_/*N*_*HD*_,
- *γ*_*HU*_ is the daily individual transition rate from *H* to *U*, and *γ*_*HU*_ = *p*_*HU*_ /*N*_*HU*_,
- *γ*_*UR*_ is the daily individual transition rate from *U* to *R*, and *γ*_*UR*_ = (1 − *p*_*UD*_)/*N*_*UR*_,
- *γ*_*UD*_ is the daily individual transition rate from *U* to *D*, and *γ*_*UD*_ = *p*_*UD*_/*N*_*UD*_,

with

- *p*_*a*_: the probability of having light symptoms or no symptoms for the infected individuals; *p*_*H*_ : the probability of needing hospitalization for mild or severely ill people; *p*_*U*_ : the probability of needing intensive care for mild or severely ill people; *p*_*HU*_ : the probability of needing intensive care under hospitalization without intensive care; *p*_*HD*_: the probability of death under hospitalization without intensive care; *p*_*UD*_: the probability of death under intensive care;
- *N*_*a*_: the number of days it takes for an asymptomatic case needs to recover; *N*_*s*_: the number of days it takes for a symptomatic case to recover without hospitalization; *N*_*IH*_ : the number of days a severely symptomatic case requires until hospitalization; *N*_*HD*_: the number of days before death in the event of hospitalization; *N*_*HU*_ : the number of days required for a hospitalized case until intensive care is provided; *N*_*UD*_: the number of days before death in the event of intensive care; *N*_*HR*_: the number of days it takes for a hospitalized case to recover; *N*_*UR*_: the number of days it takes for a case under intensive care to recover.

The infection transmission rate *β*(*t*) is the rate of the pandemic transmission from an undetected infected person to susceptible individuals at time instant *t*. As in [12], in order to combine the complex effects of lockdown strategy, a time-dependent exponentially decreasing function can be used to model the transmission rate *β*(*t*),

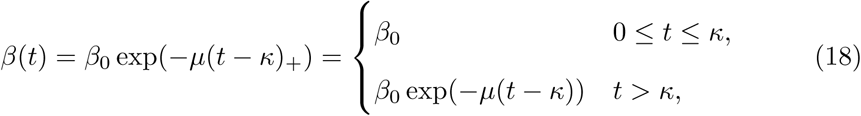

with constant parameters *β*_0_, *µ* and *κ*. Note that *β*(*t*) is constant during the initial stage of implementing effective lockdown strategies such as social distance, quarantine, healthcare, and mask worn. The transmission rate exponentially decreases at rate *µ* after these lockdown strategies take effect. The transmission rate *β*(*t*) can be illustrated in Figure 2.

**Figure 2:**
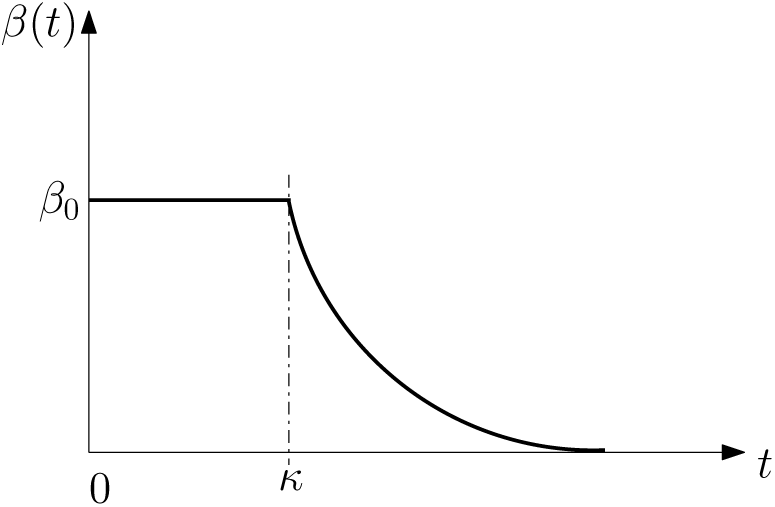
Time-evolution of the transmission rate *β* before and during the lockdown.

As one of the most critical epidemiological parameters, the basic reproductive ratio *R*_0_ defines the average number of secondary cases an average primary case produces in a totally susceptible population (see [13]). As for the model in [10], for the considered model in this paper, only the *I*^−^ individuals transmit the disease to the susceptible individuals during the early phase of outbreak. If *R*_0_ < 1 (i.e., 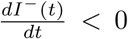), the infection “dies out” over time; inversely, if *R*_0_ > 1 (i.e., 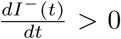), the initial number of susceptible individuals exceeds the critical threshold to allow the pandemic to spread. Thus the initial basic reproductive rate is

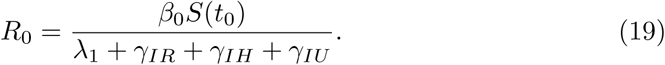

When the transmission rate *β*(*t*) and *S*(*t*) evolve as time goes by, one dynamic reproductive rate that depends on time is introduced and known as effective reproduction number *R*(*t*) in [14]. In this model, it is defined as, for *t* ≥ 0,

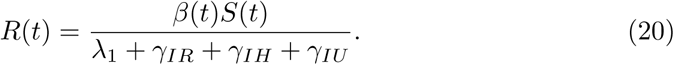

Similarly, when *R*(*t*) < 1, the number of secondary cases infected by a primary undetected infected case on day *t*, dies out over time, leading to a delay in the number of infected individuals. But when *R*(*t*) > 1, the number of undetected infected individuals grows over time. Therefore, by the control of the transmission rate *β*(*t*) that can constrain *R*(*t*) to be less than 1, the number of infected individuals grows slowly to ease the pressure on medical resources. When *S*(*t*) is bellow a threshold, the epidemic goes to extinction (see e.g., [15]). The required level of vaccination to eradicate the infection is also attained from the effective reproduction number.

The compartmental model introduced in Figure 1 exhibits a large number of unknown parameters (20 if we consider *λ*_2_ = 0). The uncertainty on these parameters can not be neglected. As an example, let us propagate uncertainty at the scale of the region *Auvergne-Rhône-Alpes*. The vector of unknown parameters is:

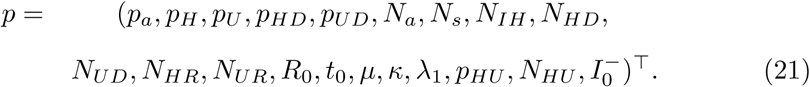

We take into account the uncertainties on these parameters by considering that each parameter is uniformly distributed with bounds consistent with typical reported values (see, e.g., [10] and references therein). Lower and upper bounds for each parameter are reported in Table 1 hereafter.

**Table 1:** Uncertainty bounds for all model parameters.

| parameters | $p_a$ | $p_H$ | $p_U$ | $p_{HD}$ | $p_{UD}$ | $N_a$ | $N_s$ | $N_{IH}$ | $N_{HD}$ | $N_{UD}$ |
| --- | --- | --- | --- | --- | --- | --- | --- | --- | --- | --- |
| lower bounds | 0.5 | 0.15 | 0.15 | 0.15 | 0.2 | 5 | 8 | 8 | 15 | 8 |
| upper bounds | 0.9 | 0.2 | 0.2 | 0.25 | 0.3 | 12 | 15 | 12 | 20 | 12 |

Table 1: Uncertainty bounds for all model parameters.
| parameters | $N_{HR}$ | $N_{UR}$ | $R_0$ | $t_0$ | $\mu$ | $\kappa$ | $\lambda_1$ | $p_{HU}$ | $N_{HU}$ | $I_0^-$ |
| --- | --- | --- | --- | --- | --- | --- | --- | --- | --- | --- |
| lower bounds | 15 | 15 | 2.5 | 2020-02-06 | 0.03 | 20 | 1e-4 | 0.001 | 1 | 1 |
| upper bounds | 25 | 20 | 4.5 | 2020-02-12 | 0.1 | 50 | 1e-3 | 0.01 | 10 | 100 |

The parameter sampling approach is based on the generation of a low-discrepancy sequence of 5000 points on the unit hypercube [0, 1]^20^. Low-discrepancy sequences have the property of uniformly and regularly filling the unit hyper-cube, without the clustering issues encountered by Monte Carlo samples. Sobol’ sequences [16] are among the best low-discrepancy sequences with solid theoretical properties and good numerical performance when dimension increases.

Figure 3 shows that the prior uncertainty is pretty high, since for example the difference between the 75 % and the 25 % quantiles for the number of people in hospital is more than 50000 at the end of the lockdown period. On Figure 4 we propagate the parameter uncertainty on the maximum number of people in intensive care units, on the date at which this maximum value is attained and on the total number of reported cases. Note that the total number of reported cases is obtained from the daily number of reported cases, *DR*, which is driven by the following equation:

**Figure 3:**
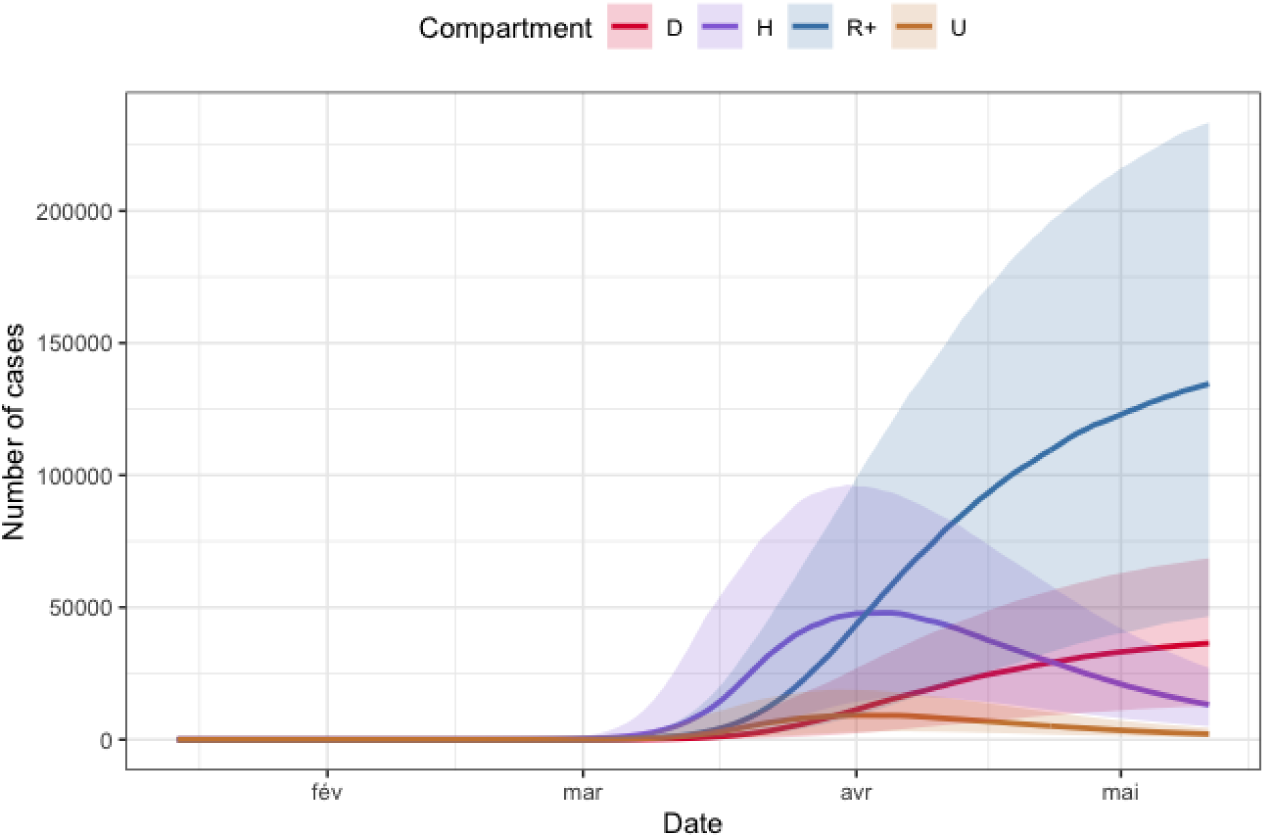
Prior uncertainty quantification for compartments *D* (in red), *H* (in purple), *R*^+^ (in blue) and *U* (in orange) for the region *Auvergne-Rhône-Alpes*. The bold lines are the pointwise medians of each functional output, whereas the colored surface is the range between the pointwise first and third quartiles.

**Figure 4:**
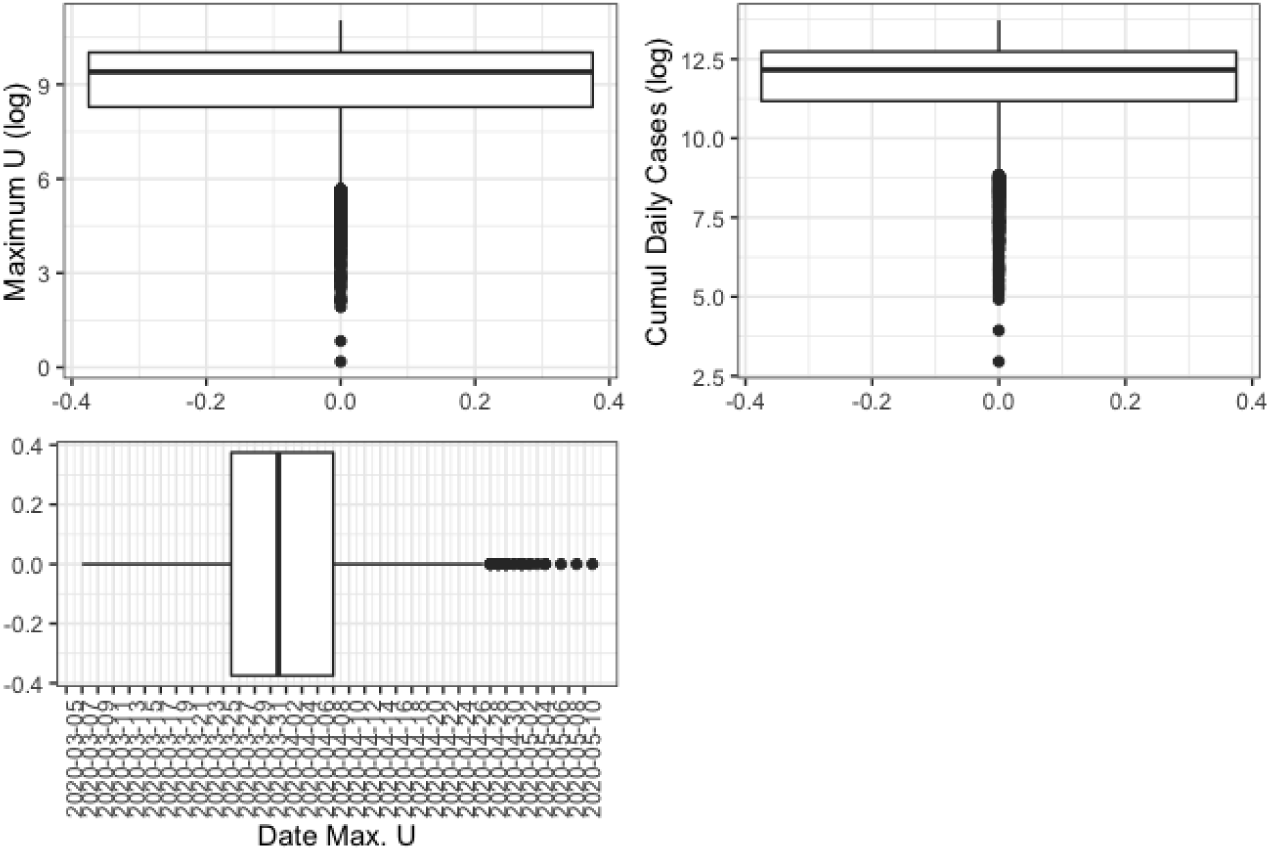
Prior uncertainty quantification for maximum value of *U* (top left, in log scale), total number of reported cases (top right, in log scale) and day where maximum value of *U* is reached for the region *Auvergne-Rhône-Alpes*.

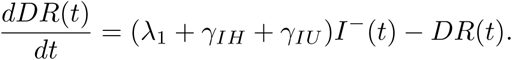

The maximum number of people in intensive care is particularly important as it provides information on the capacities the intensive care units should have to face the sanitary crisis. We show for each of these three scalar quantities of interest the boxplot which visualises five summary statistics (the median, two hinges and two whiskers) and all outlying points individually. The lower and upper hinges correspond to the first and third quartiles (the 25th and 75th percentiles). The upper whisker extends from the hinge to the largest value no further than 1.5 * IQR from the hinge (where IQR is the inter-quartile range, or distance between the first and third quartiles). The lower whisker extends from the hinge to the smallest value at most 1.5 * IQR of the hinge. Data beyond the end of the whiskers are called “outlying” points and are plotted individually. We see fpr example on these boxplots that the median for the maximum number of people in intensive care is more than 8000 with the IQR greater than 20000.

In view of the importance of uncertainties propagated from the model parameters to the quantities of interest (e.g., number of infected people at hospitals), it appears necessary to calibrate the model. Our calibration procedure is described in the next section.

## 3. Parameter identification

In this section, regional scales of France are considered and all age classes are summed to calibrate the parameters of the pandemic model (6)-(13) during confinement on the basis of data about the pandemic in France. Since all regions are not connected during the lockdown, it is sufficient to identify separtely all unknown parameters for each region. From the calibration of the model, we can observe the effects of lockdown strategies on the unknown variables, in particular for infected undetected population and recovered undetected population. The following weighted least square cost function is minimized for parameters optimization:

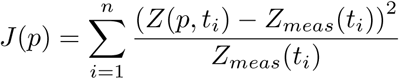

where *p* is a vector which consists of calibrated parameters; *Z*_*meas*_(*t*_*i*_) is the measured values of the corresponding observed state vector *Z*(*p, t*_*i*_) at time *t*_*i*_, *i* = 1, …, *n*, with *n* the number of days considered for calibration. This optimization problem is solved using Levenberg-Marquardt algorithm (see [17]). Since it is a local algorithm, we adopt, as in [11, Chapter 6], a multi-start approach where the initial values are obtained from a Latin Hypercube Sampling (LHS). LHS were introduced in [18] as space-filling designs on the unit hyper-cube. The LHS is built on the unit hypercube [0, 1]^20^ and then rescaled with the upper and lower bounds given in Table 1. The unknown parameter vector

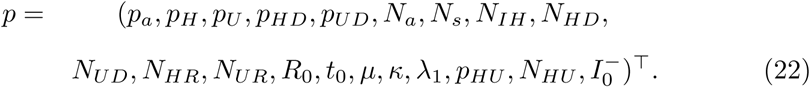

is calibrated on daily data for *H, U, D* and *R*^+^ on the lockdown period 2020-03-18 to 2020-05-11 from two data sources: the first one is a public and governmental data source [19] and the second one is a dedicated national platform with a privileged access [20].

The time step is chosen as ten percentage of one day for the numerical discretization. A general solver for ordinary differential equations is used to compute *H, U, D* or *R*^+^ for each region for all time until the end of lockdown. The results of the calibration are given in Tables 2 to 5. The results of parameter calibration for the 13 regions in France are shown in Figures 5 and 6.

**Table 2:** Optimal values of parameters *p*_*a*_, *p*_*H*_, *p*_*U*_, *p*_*HD*_, *p*_*UD*_, *N*_*a*_, *N*_*s*_ for each region.

| Regions \ Parameters | $p_a$ | $p_H$ | $p_U$ | $p_{HD}$ | $p_{UD}$ | $N_a$ | $N_s$ |
| --- | --- | --- | --- | --- | --- | --- | --- |
| Île-de-France | 0.9 | 0.15 | 0.2 | 0.20 | 0.3 | 12.0 | 13.104 |
| Centre-Val de Loire | 0.9 | 0.2 | 0.2 | 0.21 | 0.3 | 9.674 | 8.0 |
| Bourgogne-Franche-Comté | 0.83 | 0.18 | 0.2 | 0.25 | 0.3 | 6.423 | 8.576 |
| Normandie | 0.9 | 0.2 | 0.2 | 0.25 | 0.3 | 6.527 | 15.0 |
| Hauts-de-France | 0.9 | 0.15 | 0.2 | 0.244 | 0.3 | 12.0 | 15.0 |
| Grand Est | 0.9 | 0.2 | 0.2 | 0.25 | 0.3 | 12.0 | 14.551 |
| Pays de la Loire | 0.9 | 0.189 | 0.2 | 0.193 | 0.3 | 7.608 | 9.191 |
| Bretagne | 0.9 | 0.2 | 0.2 | 0.151 | 0.3 | 12.0 | 8.153 |
| Nouvelle-Aquitaine | 0.9 | 0.150 | 0.2 | 0.15 | 0.3 | 6.883 | 15.0 |
| Occitanie | 0.9 | 0.15 | 0.2 | 0.15 | 0.3 | 6.803 | 9.767 |
| Auvergne-Rhône-Alpes | 0.9 | 0.2 | 0.2 | 0.176 | 0.3 | 7.820 | 15.0 |
| Provence-Alpes-Côte d'Azur | 0.852 | 0.192 | 0.192 | 0.15 | 0.2 | 9.446 | 15.0 |
| Corse | 0.9 | 0.15 | 0.179 | 0.222 | 0.3 | 6.433 | 13.62 |

**Figure 5:**
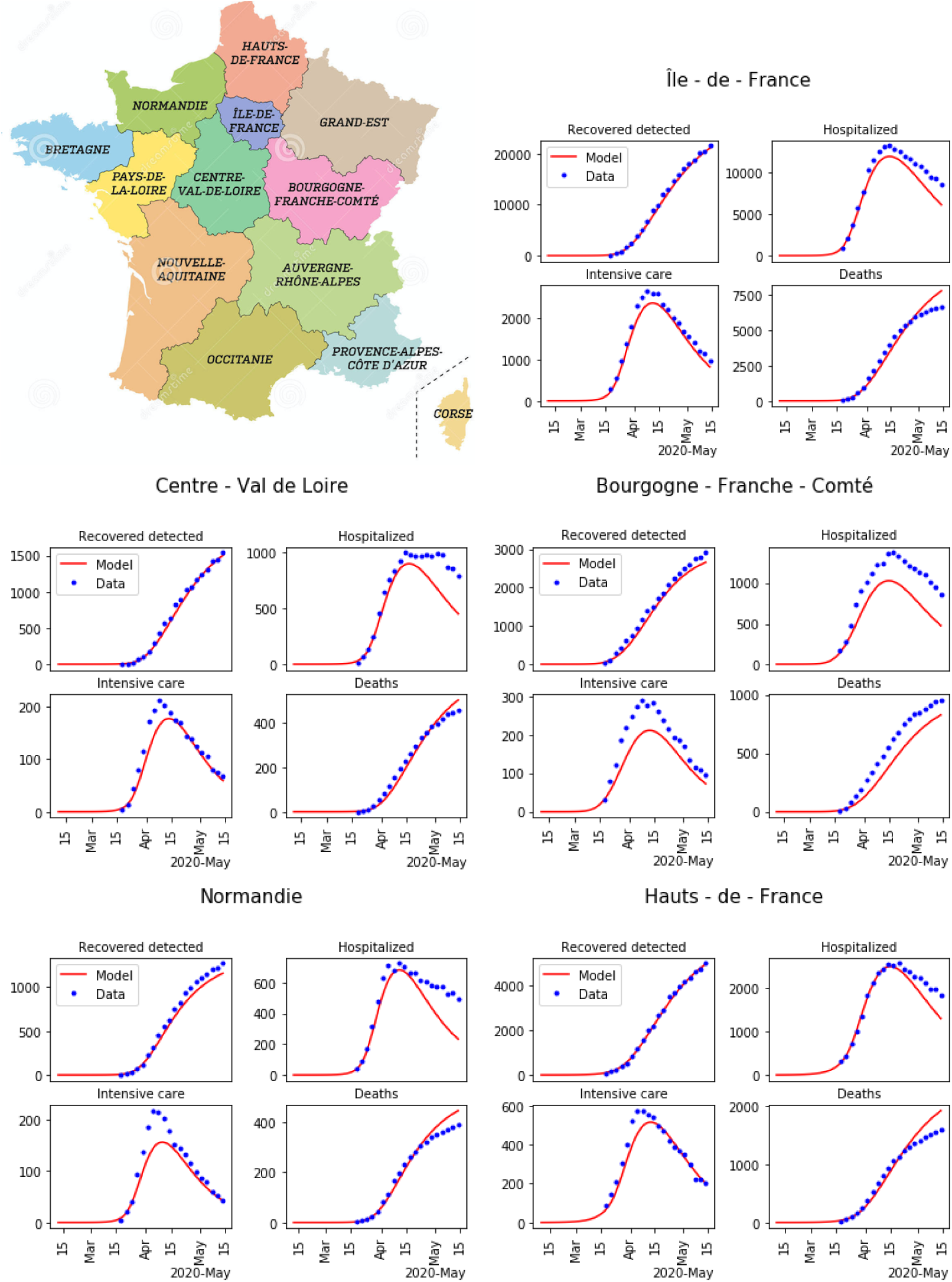
Minimap of regions in France, and result of the parameters calibration for the first 5 regions among 13 (blue dots: data, and red lines: model).

**Figure 6:**
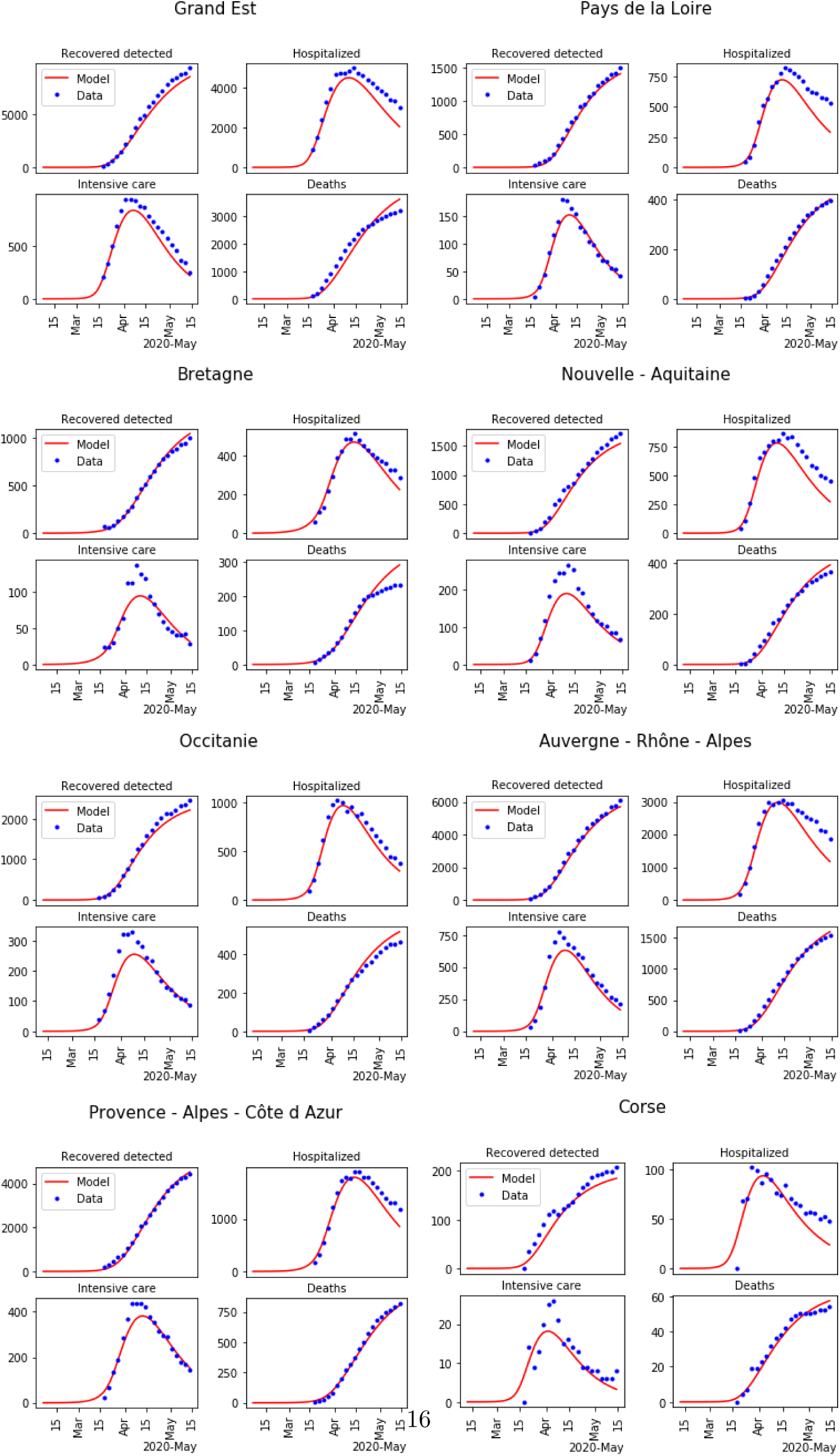
Minimap of regions in France, and result of the parameters calibration for the last 8 regions among 13 (blue dots: data, and red lines: model).

## 4. Network simulation

In order to characterize the dynamics of the pandemic transmission processes during the confinement, the epidemiological model (6)-(13) was described in the previous section. We now consider the government action of unlockdown after confinement, there is a pandemic transmission effect between each region in France. Considering *N*_*a*_ age groups, the following pandemic network model of

*N* regions is introduced, for all *j* = 1, …, *N*_*a*_ and *i* = 1, …, *N*

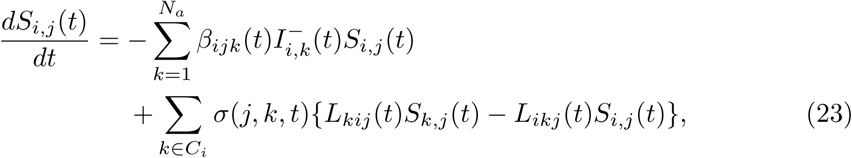

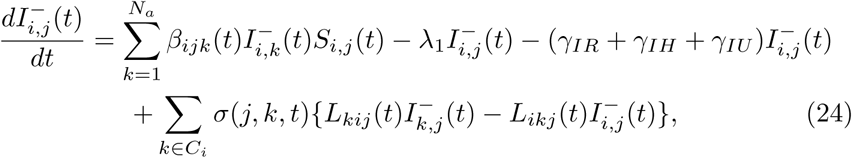

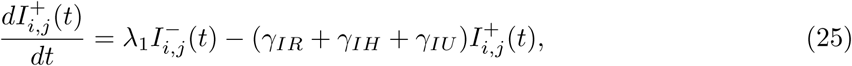

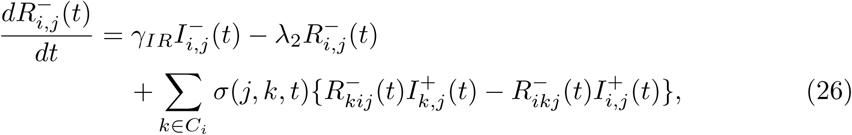

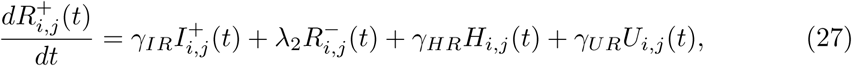

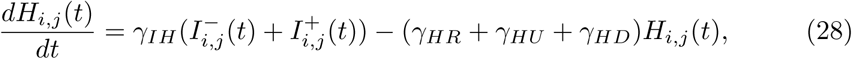

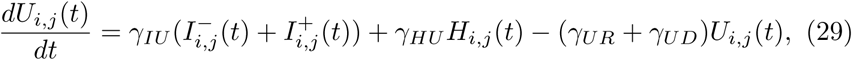

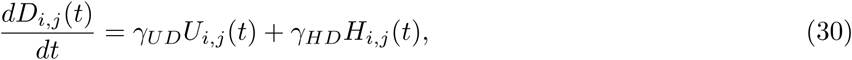

where transmission rate *β*_*ijk*_(*t*) depends only on (*i, j*), and is piecewise continuous depending on the scenario: lockdown or no-lockdown, for all *t*; for age group *j, L*_*kij*_(*t*) is the proportion of individuals moving from region *k* to region *i* in the age class *j*; the other parameters depend on the location, and also on the age group *j*; *σ*(*j, k, t*) is periodic (space dependent period *T*_*j,k*_), satisfies 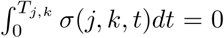, and takes value in the interval [−1, 1]; *C*_*i*_ is the set of all regions that have pandemic transmission with region *i*.

As the fast periodic switching policy in [21], we consider the inverse of the (same) exponential function of infection transmission rate *β*(*t*) in (18) to denote *β*_*ijk*_(*t*). Even though the end of confinement, the social strategies still go on, so a continuous function *β*(*t*) is used for the whole transmission process of COVID-19 from the start date of infection,

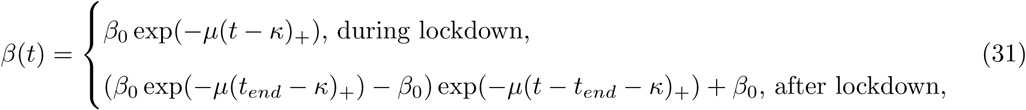

with the end time of lockdown *t*_*end*_.

The transmission rate *β*(*t*) for the whole transmission process is illustrated in Figure 7.

**Figure 7:**
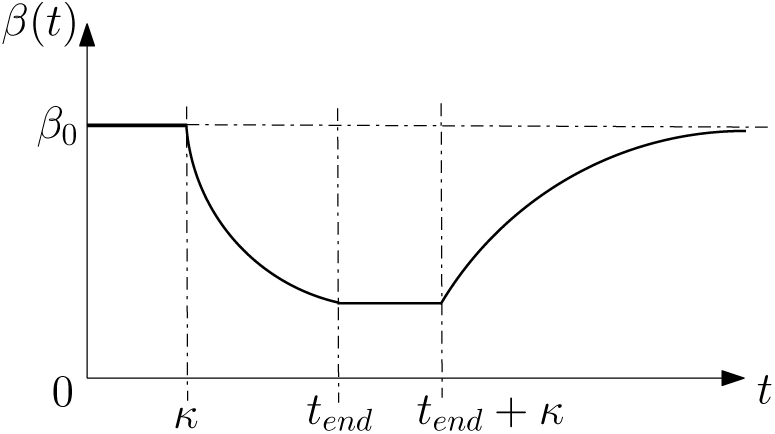
The transmission rate *β*(*t*) for the network model.

For the mobility analysis after lockdown, the mobility matrix *L* = {*L*_*ki*_}_*N*×*N*_ is computed using data of the displacement of population in France as measured by the *Institut national de la statistique et des études économiques* (INSEE). To be more specific, the professional displacements and the scholar displacements are given for each French city in [22] for some age classes. These information allow us to compute the mobility matrix *L*. The components of the matrix *L* are shown in Tables 6-8.

**Table 3:** Optimal values of parameters *N*_*IH*_, *N*_*HD*_, *N*_*UD*_, *N*_*HR*_, *N*_*UR*_, *µ* for each region.

| Parameters<br>Regions | $N_{IH}$ | $N_{HD}$ | $N_{UD}$ | $N_{HR}$ | $N_{UR}$ | $\mu$ |
| --- | --- | --- | --- | --- | --- | --- |
| Île-de-France | 12.0 | 20.0 | 9.724 | 25.0 | 20.0 | 0.10 |
| Centre-Val de Loire | 12.0 | 20.0 | 12.0 | 25.0 | 15.0 | 0.1 |
| Bourgogne-Franche-Comté | 12.0 | 20.0 | 8.391 | 25.0 | 20.0 | 0.043 |
| Normandie | 12.0 | 20.0 | 12.0 | 23.027 | 20.0 | 0.1 |
| Hauts-de-France | 12.0 | 20.0 | 9.303 | 24.331 | 20.0 | 0.1 |
| Grand Est | 11.221 | 20.0 | 8.0 | 25.0 | 20.0 | 0.0997 |
| Pays de la Loire | 12.0 | 20.0 | 12.0 | 25.0 | 20.0 | 0.1 |
| Bretagne | 12.0 | 15.225 | 12.0 | 25.0 | 15.096 | 0.1 |
| Nouvelle-Aquitaine | 12.0 | 20.0 | 12.0 | 25.0 | 20.0 | 0.1 |
| Occitanie | 12.0 | 20.0 | 12.0 | 25.0 | 20.0 | 0.1 |
| Auvergne-Rhône-Alpes | 12.0 | 20.0 | 12.0 | 25.00 | 20.0 | 0.1 |
| Provence-Alpes-Côte d'Azur | 8.867 | 20.0 | 12.0 | 25.0 | 20.0 | 0.084 |
| Corse | 12.0 | 20.0 | 12.0 | 25.0 | 20.0 | 0.1 |

**Table 4:** Optimal values of parameters *κ, λ*_1_, *p*_*HU*_, *N*_*HU*_ for each region.

| Regions \ Parameters | $\kappa$ | $\lambda_1$ | $p_{HU}$ | $N_{HU}$ |
| --- | --- | --- | --- | --- |
| Île-de-France | 35.759 | 0.0001 | 0.001 | 3.071 |
| Centre-Val de Loire | 40.141 | 0.0001 | 0.01 | 10.0 |
| Bourgogne-Franche-Comté | 25.360 | 0.001 | 0.00291 | 1.564 |
| Normandie | 37.718 | 0.000152 | 0.01 | 10.0 |
| Hauts-de-France | 40.928 | 0.000193 | 0.01 | 10.0 |
| Grand Est | 33.745 | 0.000261 | 0.0011 | 4.588 |
| Pays de la Loire | 43.524 | 0.000296 | 0.001 | 8.917 |
| Bretagne | 44.838 | 0.000259 | 0.01 | 10.0 |
| Nouvelle-Aquitaine | 36.702 | 0.000194 | 0.0064 | 2.277 |
| Occitanie | 36.056 | 0.000330 | 0.01 | 2.094 |
| Auvergne-Rhône-Alpes | 37.142 | 0.000247 | 0.001 | 3.238 |
| Provence-Alpes-Côte d'Azur | 42.813 | 0.001 | 0.01 | 10.0 |
| Corse | 29.198 | 0.000271 | 0.01 | 10.0 |

**Table 5:** Optimal values of initial conditions 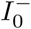, start time of infection *t*_0_ and basic reproduction rate *R*_0_.

| Regions \ Parameters | $I_0^-$ | $t_0$ | $R_0$ |
| --- | --- | --- | --- |
| Île-de-France | 30.454 | 11/02/2020 | 4.120 |
| Centre-Val de Loire | 1.025 | 11/02/2020 | 3.347 |
| Bourgogne-Franche-Comté | 1.0 | 11/02/2020 | 3.008 |
| Normandie | 1.131 | 12/02/2020 | 2.774 |
| Hauts-de-France | 200.0 | 10/02/2020 | 2.883 |
| Grand Est | 3.508 | 08/02/2020 | 4.5 |
| Pays de la Loire | 1.022 | 06/02/2020 | 2.772 |
| Bretagne | 50.582 | 06/02/2020 | 2.5 |
| Nouvelle-Aquitaine | 1.282 | 11/02/2020 | 2.950 |
| Occitanie | 14.708 | 12/02/2020 | 2.548 |
| Auvergne-Rhône-Alpes | 15.620 | 11/02/2020 | 2.884 |
| Provence-Alpes-Côte d'Azur | 25.299 | 06/02/2020 | 2.583 |
| Corse | 1.0 | 12/02/2020 | 2.859 |

**Table 6:** First part of components *L*_*ki*_ in the mobility matrix *L*.

| Region i \ Region k | Île-de-France | Centre-Val de Loire | Bourgogne-Franche-Comté | Normandie |
| --- | --- | --- | --- | --- |
| Île-de-France | 0.000e+00 | 3.662e-04 | 1.870e-04 | 3.828e-04 |
| Centre-Val de Loire | 5.485e-03 | 0.000e+00 | 8.279e-04 | 5.642e-04 |
| Bourgogne-Franche-Comté | 1.136e-03 | 4.346e-04 | 0.000e+00 | 5.651e-05 |
| Normandie | 2.803e-03 | 6.993e-04 | 5.574e-05 | 0.000e+00 |
| Hauts-de-France | 4.077e-03 | 6.803e-05 | 8.013e-05 | 5.169e-04 |
| Grand Est | 7.146e-04 | 5.116e-05 | 5.833e-04 | 6.038e-05 |
| Pays de la Loire | 7.411e-04 | 6.476e-04 | 5.488e-05 | 8.895e-04 |
| Bretagne | 6.099e-04 | 1.103e-04 | 4.689e-05 | 4.358e-04 |
| Nouvelle-Aquitaine | 6.153e-04 | 3.345e-04 | 6.910e-05 | 7.362e-05 |
| Occitanie | 4.616e-04 | 5.165e-05 | 7.992e-05 | 5.219e-05 |
| Auvergne-Rhône-Alpes | 4.855e-04 | 1.066e-04 | 8.104e-04 | 5.503e-05 |
| Provence-Alpes-Côte d'Azur | 5.062e-04 | 4.511e-05 | 8.287e-05 | 5.736e-05 |
| Corse | 8.718e-04 | 1.388e-04 | 1.388e-04 | 9.041e-05 |

**Table 7:**
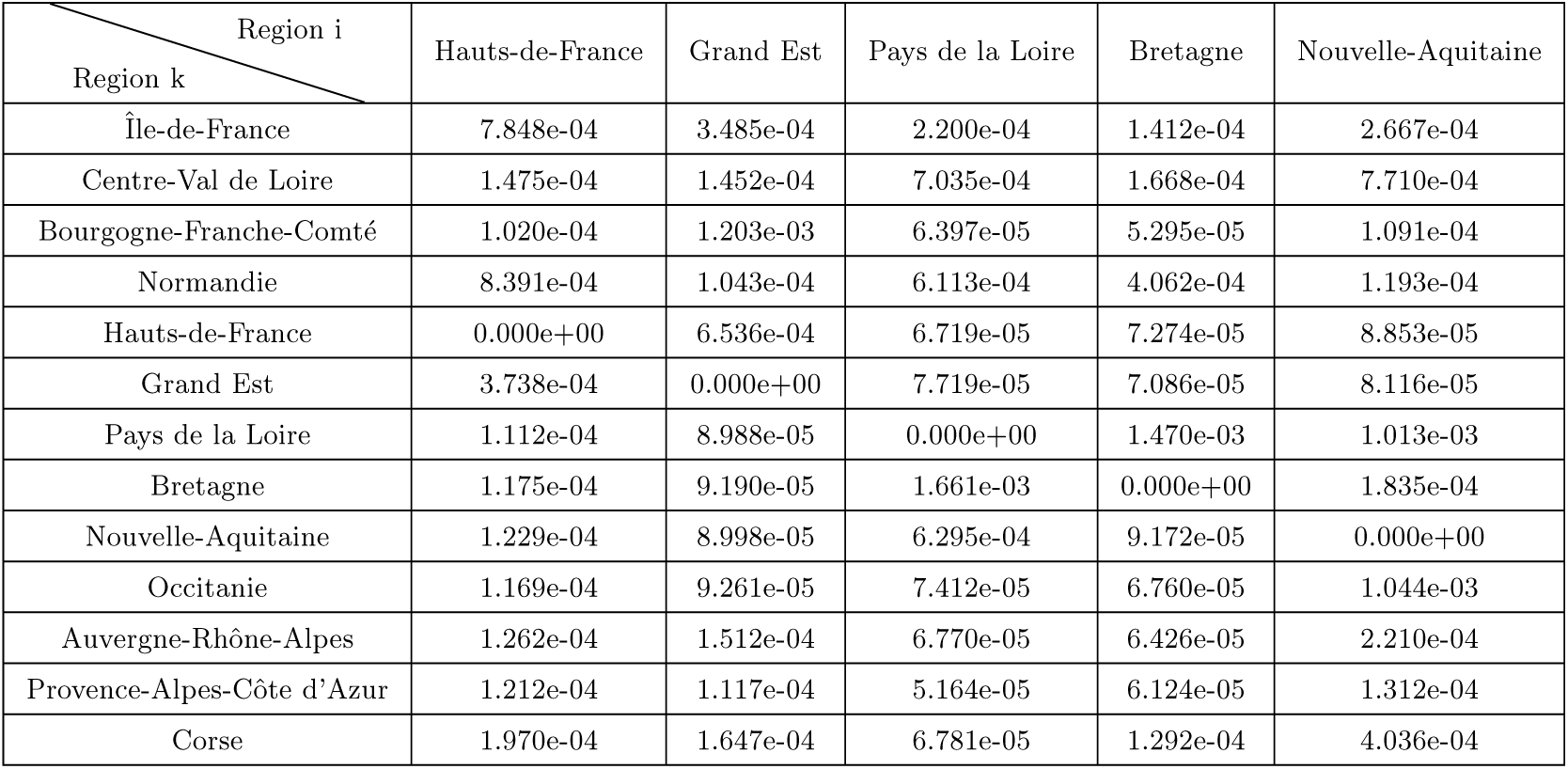
Second part of components *L*_*ki*_ in the mobility matrix *L*.

| Region i<br>Region k | Hauts-de-France | Grand Est | Pays de la Loire | Bretagne | Nouvelle-Aquitaine |
| --- | --- | --- | --- | --- | --- |
| Île-de-France | 7.848e-04 | 3.485e-04 | 2.200e-04 | 1.412e-04 | 2.667e-04 |
| Centre-Val de Loire | 1.475e-04 | 1.452e-04 | 7.035e-04 | 1.668e-04 | 7.710e-04 |
| Bourgogne-Franche-Comté | 1.020e-04 | 1.203e-03 | 6.397e-05 | 5.295e-05 | 1.091e-04 |
| Normandie | 8.391e-04 | 1.043e-04 | 6.113e-04 | 4.062e-04 | 1.193e-04 |
| Hauts-de-France | 0.000e+00 | 6.536e-04 | 6.719e-05 | 7.274e-05 | 8.853e-05 |
| Grand Est | 3.738e-04 | 0.000e+00 | 7.719e-05 | 7.086e-05 | 8.116e-05 |
| Pays de la Loire | 1.112e-04 | 8.988e-05 | 0.000e+00 | 1.470e-03 | 1.013e-03 |
| Bretagne | 1.175e-04 | 9.190e-05 | 1.661e-03 | 0.000e+00 | 1.835e-04 |
| Nouvelle-Aquitaine | 1.229e-04 | 8.998e-05 | 6.295e-04 | 9.172e-05 | 0.000e+00 |
| Occitanie | 1.169e-04 | 9.261e-05 | 7.412e-05 | 6.760e-05 | 1.044e-03 |
| Auvergne-Rhône-Alpes | 1.262e-04 | 1.512e-04 | 6.770e-05 | 6.426e-05 | 2.210e-04 |
| Provence-Alpes-Côte d'Azur | 1.212e-04 | 1.117e-04 | 5.164e-05 | 6.124e-05 | 1.312e-04 |
| Corse | 1.970e-04 | 1.647e-04 | 6.781e-05 | 1.292e-04 | 4.036e-04 |

**Table 8:** Third part of components *L*_*ki*_ in the mobility matrix *L*.

| Region i<br>Region k | Occitanie | Auvergne-Rhône-Alpes | Provence-Alpes-Côte d'Azur | Corse |
| --- | --- | --- | --- | --- |
| Île-de-France | 2.133e-04 | 3.019e-04 | 1.429e-04 | 1.001e-05 |
| Centre-Val de Loire | 1.209e-04 | 3.582e-04 | 6.160e-05 | 2.747e-06 |
| Bourgogne-Franche-Comté | 1.009e-04 | 1.918e-03 | 1.084e-04 | 3.198e-06 |
| Normandie | 8.647e-05 | 1.681e-04 | 6.719e-05 | 2.712e-06 |
| Hauts-de-France | 8.618e-05 | 1.409e-04 | 5.090e-05 | 1.008e-06 |
| Grand Est | 7.556e-05 | 1.791e-04 | 1.106e-04 | 1.808e-06 |
| Pays de la Loire | 9.884e-05 | 2.159e-04 | 5.404e-05 | 2.520e-06 |
| Bretagne | 1.097e-04 | 1.638e-04 | 6.189e-05 | 6.252e-06 |
| Nouvelle-Aquitaine | 9.997e-04 | 3.356e-04 | 9.190e-05 | 4.873e-06 |
| Occitanie | 0.000e+00 | 6.060e-04 | 1.626e-03 | 7.430e-06 |
| Auvergne-Rhône-Alpes | 3.787e-04 | 0.000e+00 | 4.293e-04 | 6.202e-06 |
| Provence-Alpes-Côte d'Azur | 1.115e-03 | 8.193e-04 | 0.000e+00 | 2.613e-05 |
| Corse | 5.199e-04 | 3.487e-04 | 1.698e-03 | 0.000e+00 |

We choose the same time step as for the calibration step. The simulation results of the considered network model for 13 regions in France are shown in Figures 8 and 9, all parameters and the values of all states at the starting date of lockdown have been identified during lockdown, the end date of confinement is 11th of May in France.

**Figure 8:**
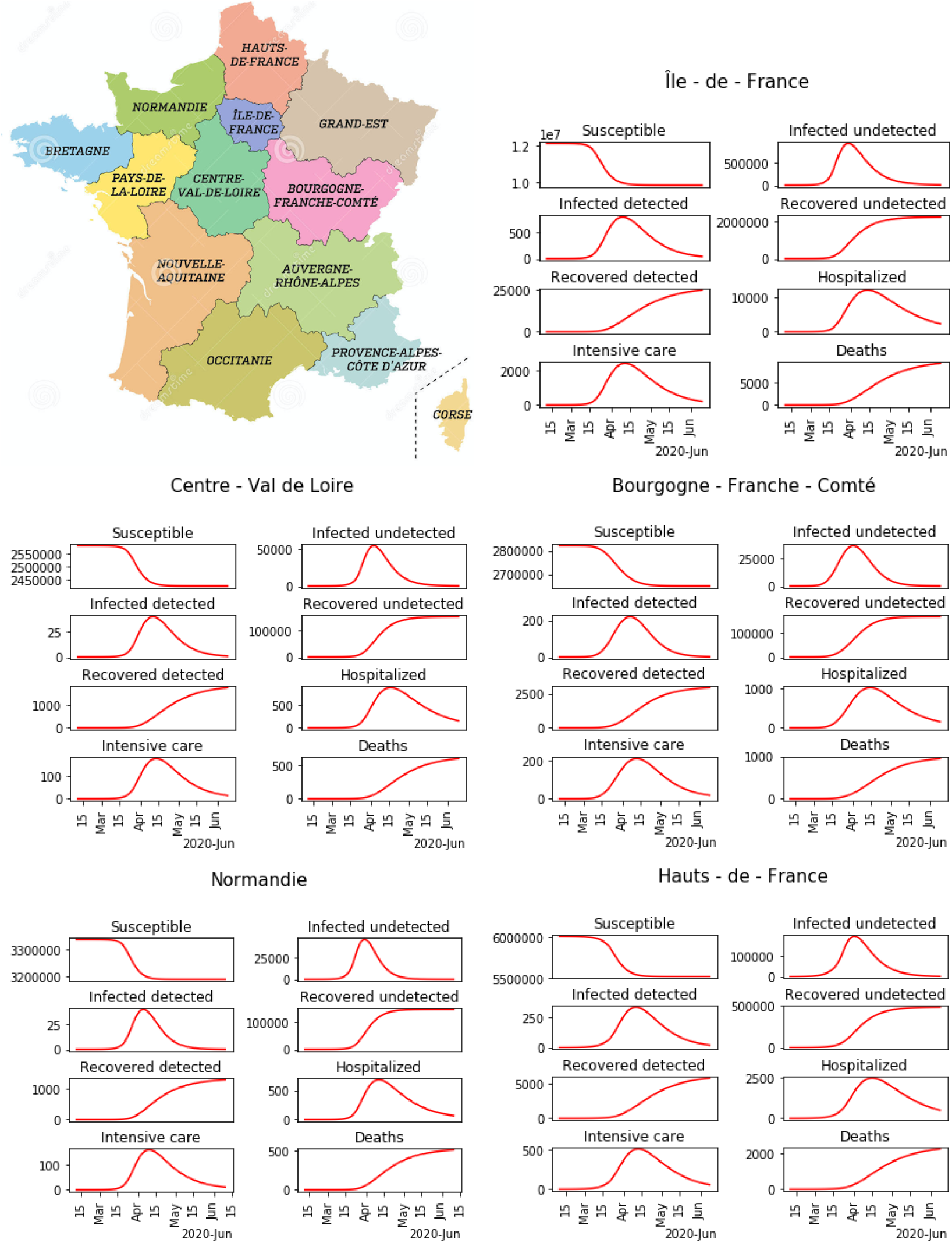
Minimap of regions in France, and the simulation of the pandemic Network model for the first 5 regions among 13.

**Figure 9:**
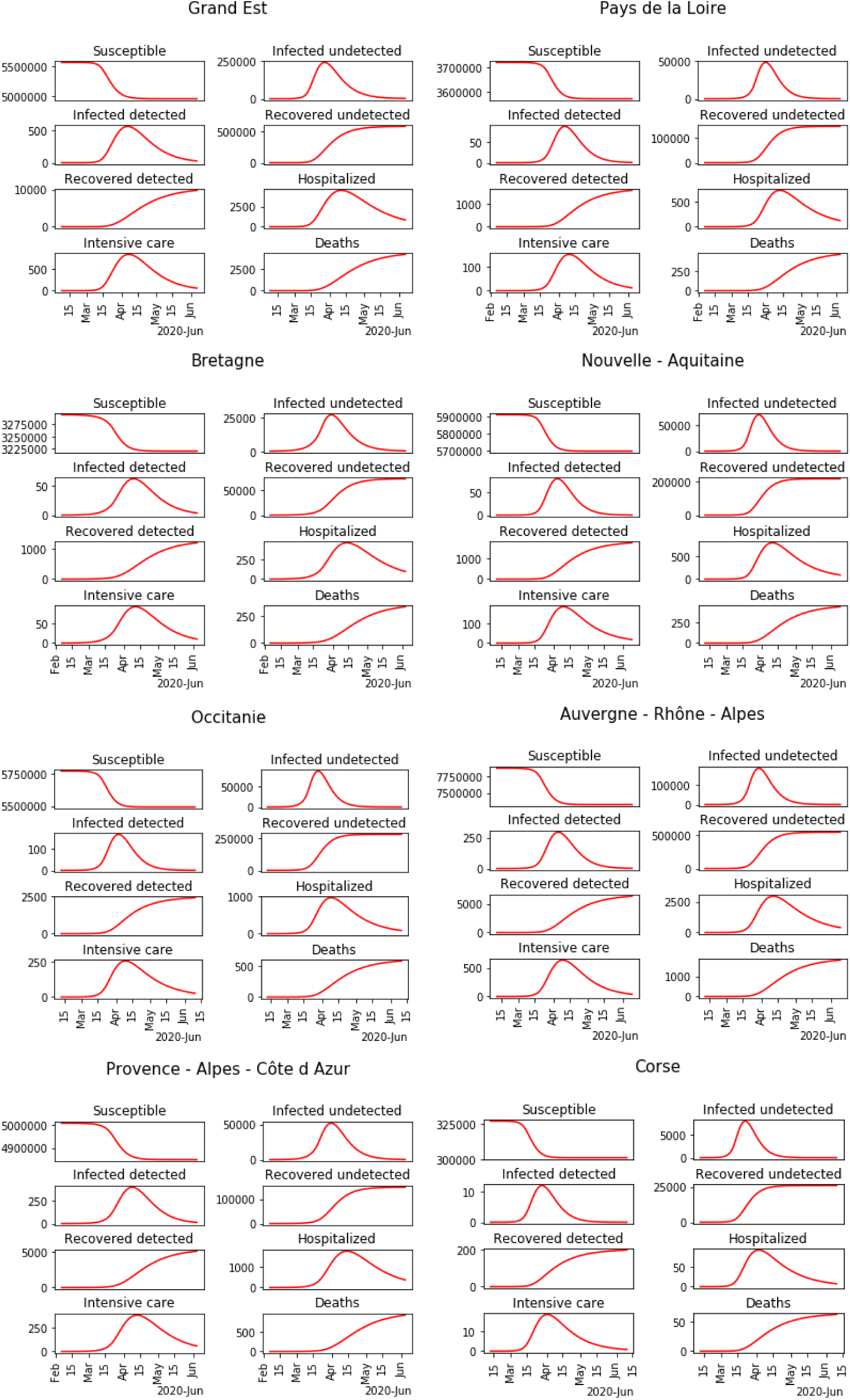
Minimap of regions in France, and the simulation of the pandemic Network model for the last 8 regions among 13.

## 5. Network of cities

In this section, we use the parameter identification method developed in Section 3 to simulate another network of areas. Instead of considering the network of metropolitan regions as in Section 4, we consider the network of all French cities. There are around 36.000 cities in France, and INSEE measures the displacement of people between each couple of cities [22]. To simulate the transport effect on the pandemic dynamics, we follow the same approach as in Section 4. To be more specific, we use the same model as (23)-(30) but instead of considering *N* = 13 regions, we consider *N* = 36.000 cities with only one age class (*j* = 1):

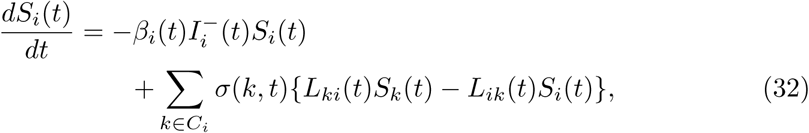

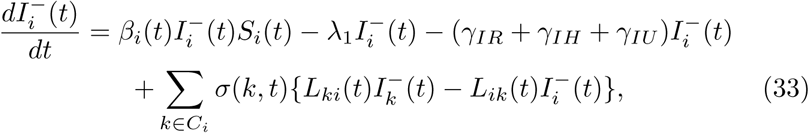

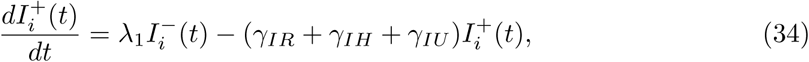

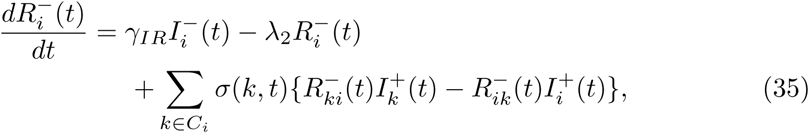

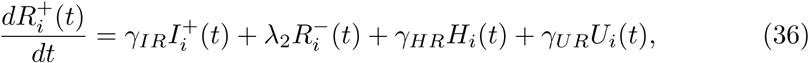

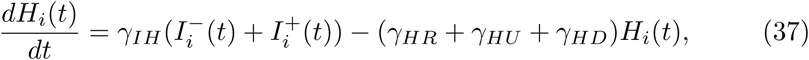

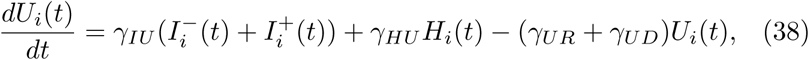

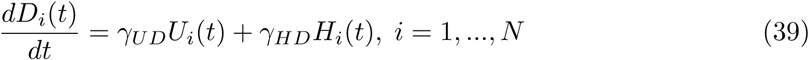

where *L*_*ki*_(*t*) is the proportion of individuals moving from city *k* to city *i*, and is derived from the real data of INSEE, and *C*_*i*_ is the set of all cities that have pandemic transmission with city *i*. All the other parameters are chosen as the ones of the region to which each city belongs.

To simulate this system of 8 ∗ 36.000 differential equations, we now specify initial conditions. To simplify, the epidemic start date of each city is taken as the same as the epidemic start date of the region to which it belongs, and the initial condition for the undetected infected individuals 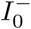 for the capital of each region is the one of the region, while it is set to 0 for all other cities in the region. It is equivalent to say that the pandemic dynamics starts at the capital of each region. The population of each French city is used as initial condition for the susceptible individuals.

The transport effect between cities is seen on Figures 10-12. On these Figures, we observe the spatial evolution of the pandemics between 2020-03-17 and 2020-08-01. At the early date, the results are impacted by the initial conditions. Indeed the infected people are mainly concentrated in the capital of each region. Then the pandemic spreads to the other cities. Note that we did not model the wearing of cloth face coverings in public settings, which could be included in the modelling of the transmission rate *β*(*t*).

**Figure 10:**
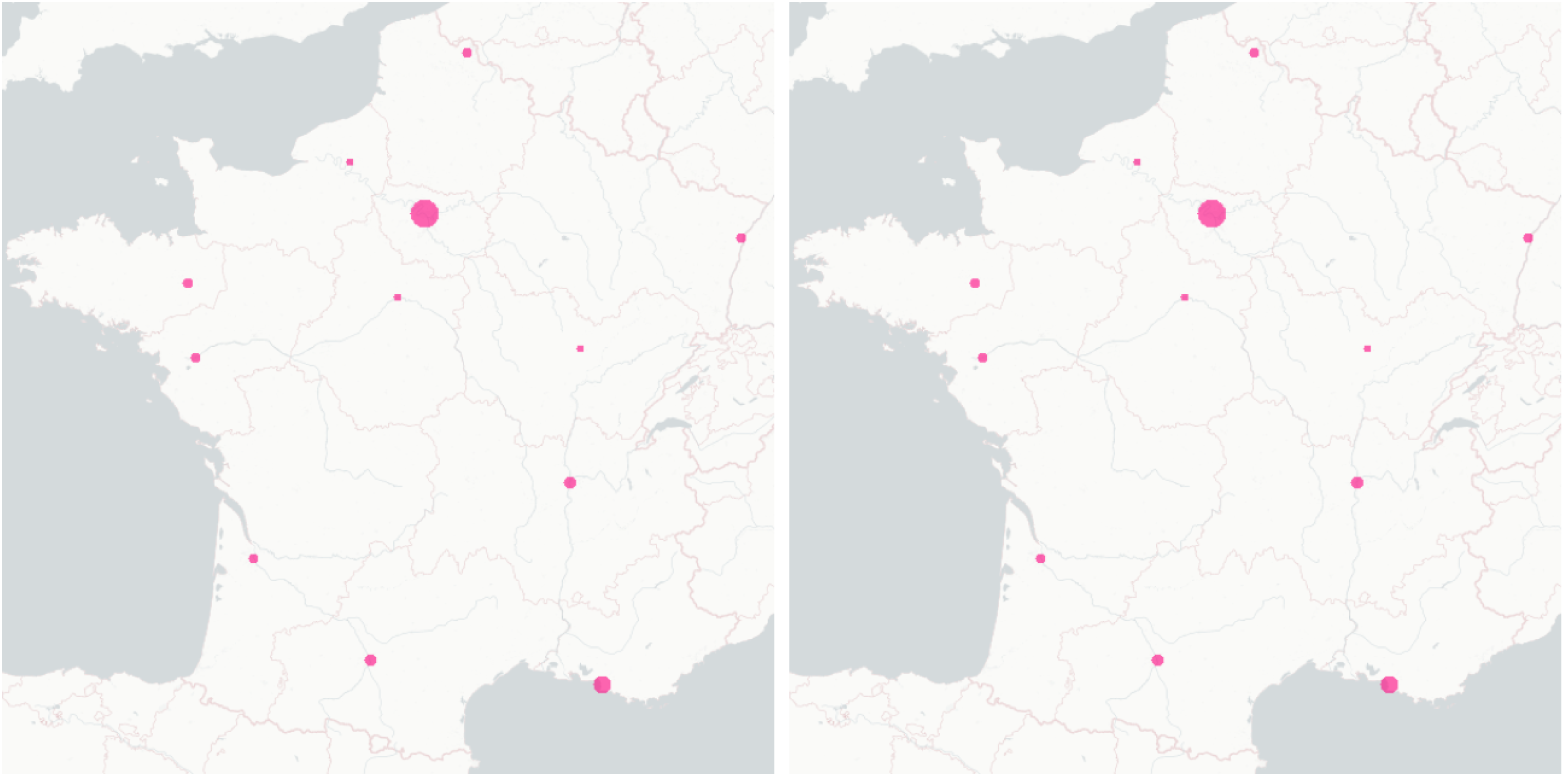
The maps of the transport effect between cities in France (undetected infected plus detected infected from 0% (blue) to 2% (magenta) of the population for each commune): the date for the map on the left is 2020-03-17 (start date of the lockdown in France) and the one for the map on the right is 2020-04-01.

**Figure 11:**
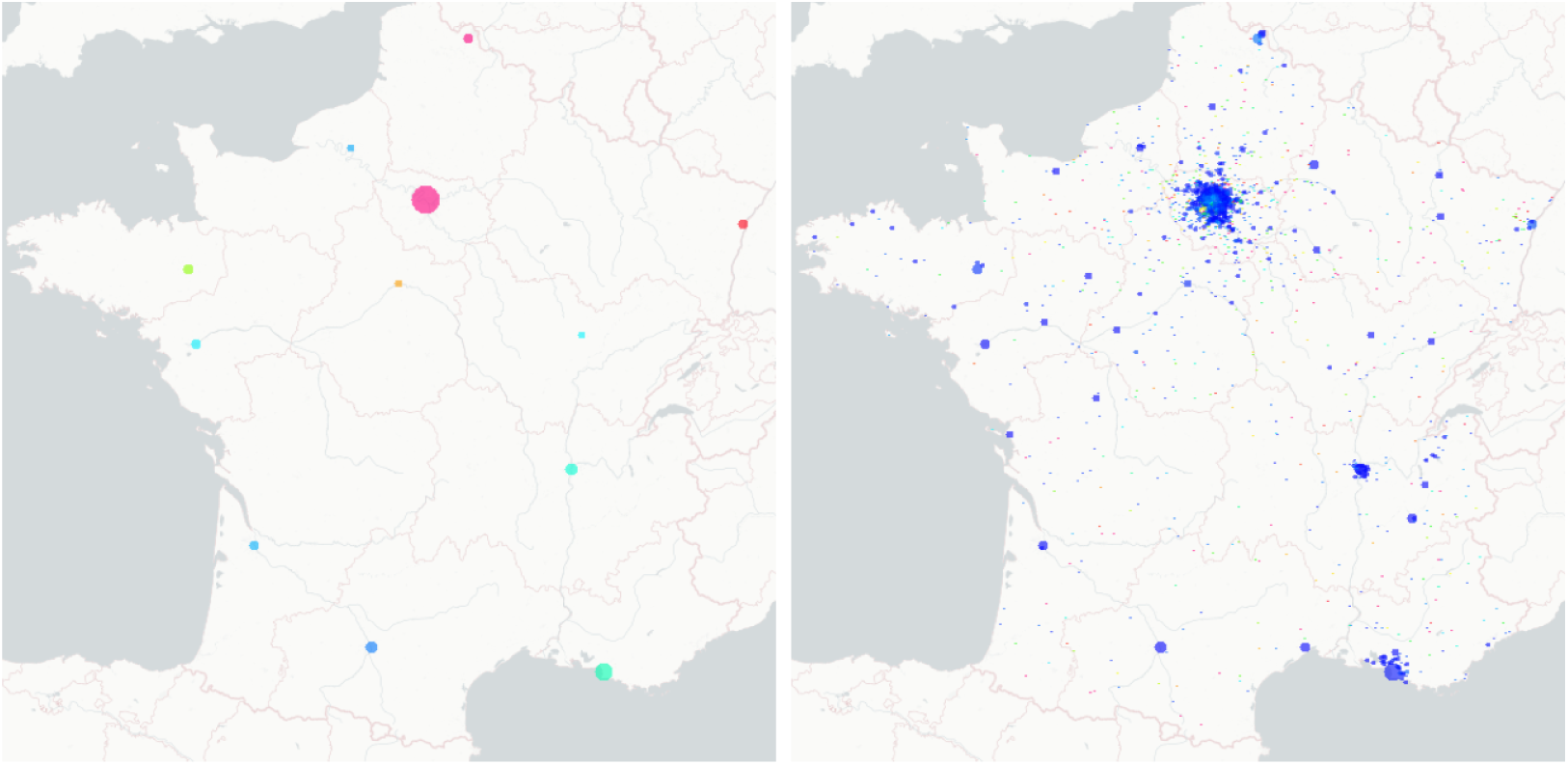
The maps of the transport effect between cities in France (undetected infected plus detected infected from 0% (blue) to 2% (magenta) of the population for each commune): the date for the map on the left is 2020-05-01 and the one for the map on the right is 2020-06-01.

**Figure 12:**
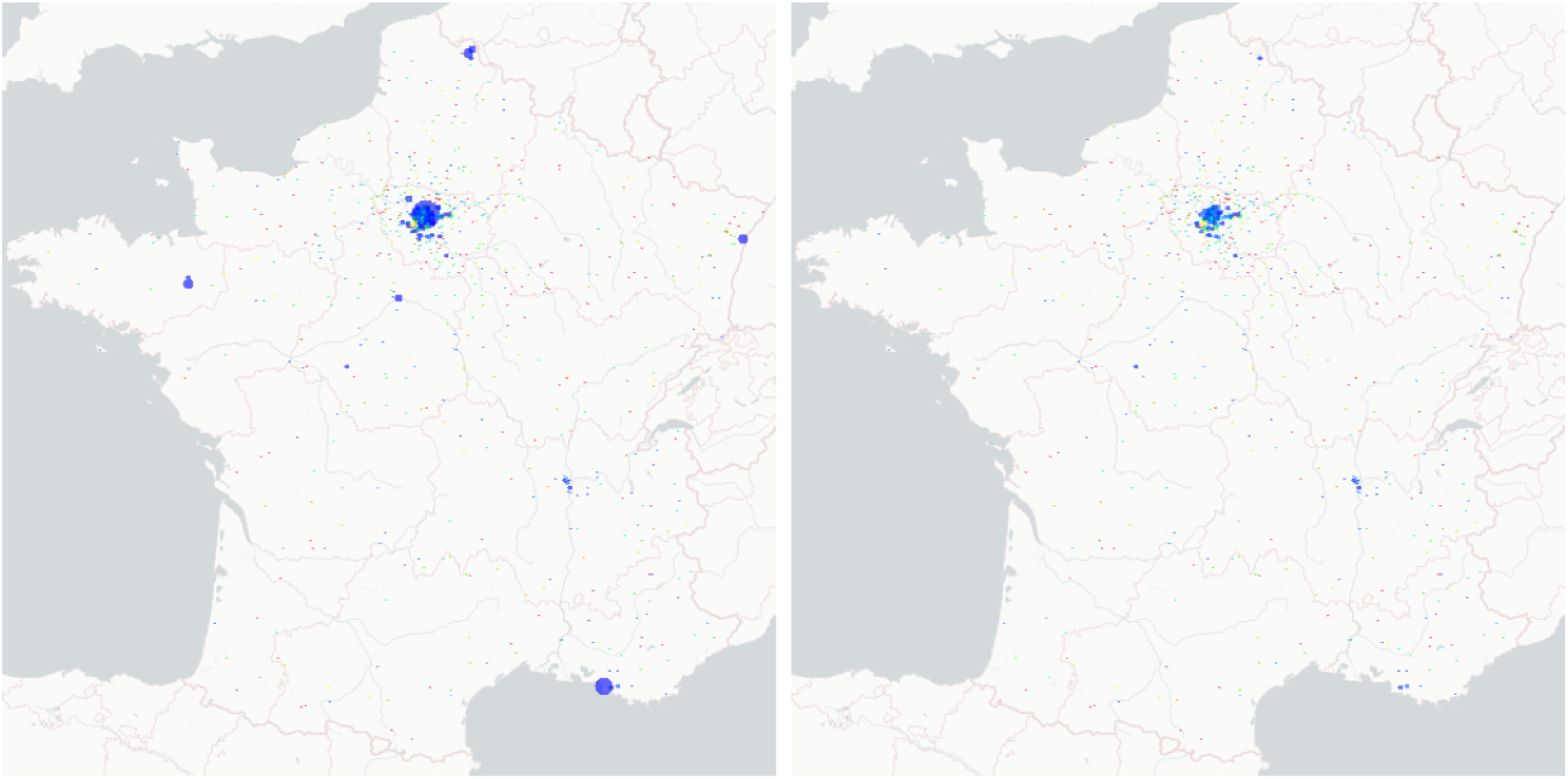
The maps of the transport effect between cities in France(undetected infected plus detected infected from 0% (blue) to 2% (magenta) of the population for each commune): the date for the map on the left is 2020-07-01 and the one for the map on the right is 2020-08-01.

## 6. Discussion and a new integro-differential model

In this section, the general form of an integro-differential model capable of integrating different age classes and areas is introduced to discuss the transport effect of COVID-19 in France after lockdown. By “areas” we mean a given geographical scale as the set of 13 Metropolitan regions (as considered in Section 4), or the set or all 101 French departments, or all cities (as considered in Section 5), or other geographical areas. For each age class *a* ∈ *ages* in area *x* ∈ *areas*, we consider the following integro-differential equations, for any time *t* ≥ 0 after confinement,

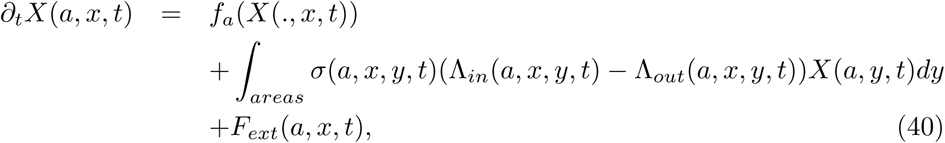

with

- *ages, N*_*a*_ ∈ ℕ, the set of different age classes of population, depending on the age scale under study. As an example, we can consider all scholar age classes, or elderly ages, or a mix of such age classes as the set

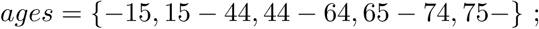
- *areas*, the set of different areas of population under study, depending on the considered geographical scale. As an example, considering all metropolitan regions, as considered in Section 4, yields the set

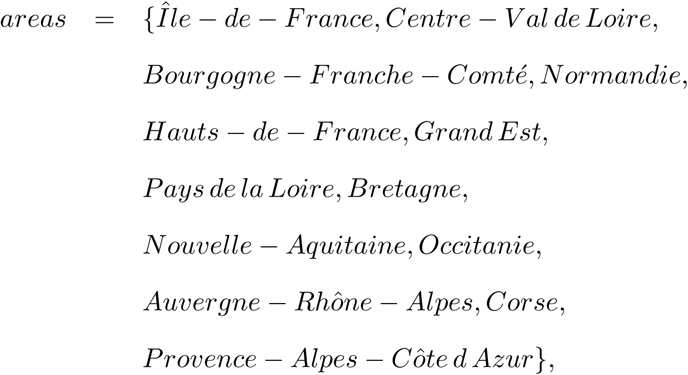 As another example, considering all French departments gives a set of 101 areas, or considering the geographical scale of French cities yield a set of around 36.000 areas, as considered in Section 5 and so on… We can even consider set of countries to model the international transport effect.
- *X*(*a, x, t*) ∈ ℝ^8^ is the 8-vector consisting of compartments of the age class *a*, in the area *x*, at time *t*;
- For all age class *a, f*_*a*_(*X*(., *x, t*)) is the pandemic transmission dynamics for age class *a* from all other age classes in the area *x* at time *t*. Without considering the age effect, it is given by the right-hand side of systems (6)-(13). Inspired by the contact matrix approach developed in e.g. [23, Chapter 3, Page 76], by considering multiple age classes, the transmission term is the following integral

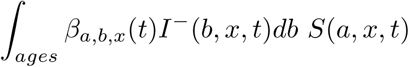

instead of

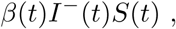

where *β*_*a,b,x*_(*t*) is the contact function between age classes *a* and *b*, in the area *x*, and at time *t*. Therefore the function *f*_*a*_ is given by

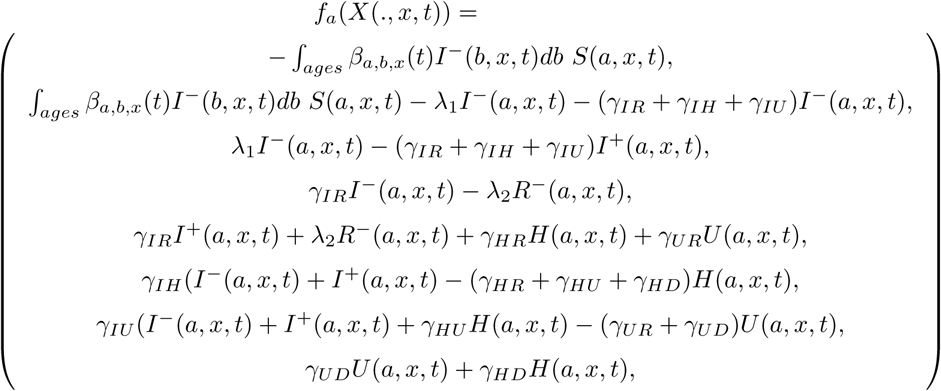

where all parameters depend on the age class *a* and the area *x*;

- Λ_*in*_(*a, x, y, t*) ∈ ℝ is the density of people coming (*in*) area *x* from area *y* ∈ *areas* at time *t*, for age class *a*;
- Λ_*out*_(*a, x, y, t*) ∈ ℝ is the density of people going to (*out*) area *y* ∈ *areas* from area *x* at time *t*, for age class *a*;
- *F*_*ext*_(*a, x, t*) ∈ ℝ^8^ is the external flux coming into location *x* at time *t* in the age class *a*. As an example for the simulations of Section 4 (where the metropolitan regions are considered) and of Section 5 (where all French cities are considered), it is 0 because the boundary of France are close (at the time of the simulation);
- *σ*(*a, x, y, t*) is the lockdown function for the age class *a*, between the areas *x* and *y* at time *t*. As an example, before the 11th of May, it was forbidden to travel for more than 100km in France. Such a policy could depend on the age classes and on the areas, e.g., to control so called “clusters” of COVID-19;
- ∫*_areas_ σ*(*a, x, y, t*)Λ_*in*_(*a, x, y, t*)*X*(*a, y, t*)*dy* provides the total number of people coming into area *x* from all the other areas.

Equation (40) describes the network dynamics of COVID-19 pandemic after lockdown and the transport effect on different age class on the basis of the regional pandemic transmission dynamics during lockdown. The proposed structure makes it easier to understand different forms of the kernel. The interest of this model is that it could be adapated to any geographical scales, and to all age classes. For a control point of view, the most important term is *σ*(*a, x, y, t*) which defines the lockdown policy that defines the mobility between areas *x* and *y* at time *t* for the age class *a*. Many control problems could be studied for this model, as optimal control to reduced the pandemic effect, or to minimize the mortality in particular. It is of great importance for the mobility dynamics of the pandemic.

Beyond that, inspired by advection-diffusion modelling of population dynamics (as considered in [24]), it is natural to model the displacement inside a given area by a diffusion term (see [25]). The corresponding model is formulated as follows:

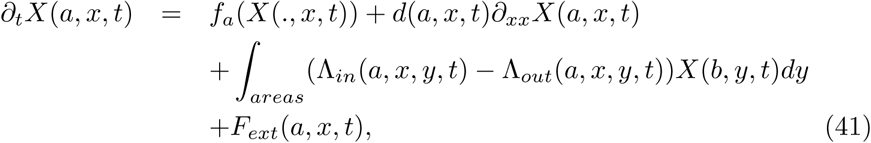

where the diffusion coefficient *d*(*a, x, t*) is a function that depends on age class *a*, areas *x* and time *t*.

This 2-order partial differential equation predicts that for age class *a* in the area *x*, how diffusion causes the number of individuals in the different compartments, especially undetected infectives and deaths, to change with respect to time *t* after lockdown. As long as one susceptible person is infected after directly or indirectly contacting disease carriers in the area *x*, diffusion takes place. When the number of infectious individuals in a local area is low compared to the surrounding areas, the pandemic will diffuse in from the surroundings, so the number of infectives in this area will increase. Conversely, the pandemic will diffuse out and the number of infectives will increase in the surrounding areas. The process of diffusion is influenced by distance, nearby individuals or areas have higher probability of contact than remote individuals or areas.

Finally, gender differentiation or other properties may be taken into account to characterize types of populations and to study the optimal lockdown control of pandemic dynamics based on our previous work. It is worth stressing that, in the long run, optimal lockdown strategies should consider the balance between the lower number of deaths and minimum healthcare and social costs.

## 7. Conclusion

In this paper, we investigated an extended model of the classical SIRD pandemic model to characterize the regional transmission of COVID-19 after lockdown in France. Incorporating the time delays arising from incubation, testing and the complex effects of government measures, an exponential function of the transmission rate *β*(*t*) was presented for the regional model. By fitting the regional model to the real data, the optimal parameters of this regional model for each region in France were derived. Based on the previous results of the extended model, we introduced and simulated a network model of pandemic transmission between regions after confinement in France while considering age classes. Regarding the transmission rate *β*(*t*) for the network model, we selected the inverse function of the previous *β*(*t*) to contribute to the transport effect after lockdown. By using the same model and method, we simulated the pandemic network for all cities in Franc to visualize the transport effects of the pandemic between cities. Considering age classes, we discussed an integro-differential equation for modelling the network of infectious diseases in the discussion part.

Because of the large volumes of data and complicated calculations needed for parameters calibration and simulation when considering many geographical areas and many age classes, the requirements in terms of computer hardware and software are rather high. In order to achieve accurate results, appropriate and efficient data processing methods will be applied. Moreover appropriate dedicated theoretical work is needed to study the integro-differential model derived in Section 6.

In future works, we will formulate and study optimal control problems in order to balance the induced sanitary and economic costs. The lockdown strategies implemented in France should be evaluated and compared to the proposed optimal strategies.

## Data Availability

Calibrate on daily data for H, U, D and R+ on the lockdown period 2020-03-18 to 2020-05-11 from two data sources: the first one is Gouvernement fran ̧cais, Info coronavirus covid 19, and the second one is Minist`ere des solidarit ́es et de la sant ́e, Plateforme COVID-19.

https://www.gouvernement.fr/info-coronavirus/carte-et-donnees

https://www.francetvinfo.fr/sante/maladie/coronavirus/coronavirus-450-personnes-en-quatorzaine-apres-des-cas-dans-une-ecole-de-lyon_4036889.html

## Acknowledgements

The authors are very greatfull to Sébastien Da Veiga, Senior Expert in Statistics and Optimization at SafranTech (France) for the R codes used for calibration and uncertainty calibration.

